# A Mechanism for Severity of Disease in Older Patients with COVID-19: The Nexus between Telomere Length and Lymphopenia

**DOI:** 10.1101/2020.10.01.20205393

**Authors:** Athanase Benetos, Tsung-Po Lai, Simon Toupance, Carlos Labat, Simon Verhulst, Sylvie Gautier, Marie-Noelle Ungeheuer, Christine Perret-Guillaume, Daniel Levy, Ezra Susser, Abraham Aviv

## Abstract

**Background:** Lymphopenia due to a plummeting T-cell count is a major feature of severe COVID-19. T-cell proliferation is telomere length (TL)-dependent and TL shortens with age. Older persons are disproportionally affected by severe COVID-19, and we hypothesized that those with short TL have less capacity to mount an adequate T-cell proliferative response to SARS-CoV-2. This hypothesis predicts that among older patients with COVID-19, shorter telomeres of peripheral blood mononuclear cells (PBMCs) will be associated with a lower lymphocyte count.

**Methods:** Our sample comprised 17 COVID-19 and 21 non-COVID-19 patients, aged 87 ± 8 (mean ± SD) and 87 ± 9 years, respectively. We measured TL by the Telomere Shortest Length Assay, a novel method that measures and tallies the short telomeres directly relevant to telomere-mediated biological processes. The primary analysis quantified TL as the proportion of telomeres shorter than 2 kilobases. For comparison, we also quantified TL by Southern blotting, which measures the mean length of telomeres.

**Results:** Lymphocyte count (10^9^/L) was 0.91 ± 0.42 in COVID-19 patients and 1.50 ± 0.50 in non-COVID-19 patients (P < 0.001). In COVID-19 patients, but not in non-COVID-19 patients, lymphocyte count was inversely correlated with the proportion of telomeres shorter than 2 kilobases (P = 0.005) and positively correlated with the mean of telomeres measured by TeSLA (P = 0.03). Lymphocyte counts showed no statistically significant correlations with Southern blotting results in COVID-19 or non-COVID-19 patients.

**Conclusions:** These results support the hypothesis that a compromised TL-dependent T-cell proliferative response contributes to lymphopenia and the resulting disproportionate severity of COVID-19 among old adults. We infer that infection with SARS-CoV-2 uncovers the limits of the TL reserves of older persons.

---

A profound drop in T-cell count is the principal cause of COVID-19 lymphopenia,^1-4^ whose magnitude is an indicator of COVID-19 severity. ^5,6^ The drop in T cells reflects a failure of T-cell proliferation to keep up with the drastic T-cell loss from the circulation during the course of the illness. Greatly accelerated T-cell proliferation is essential, therefore, to offset the development of and steer the recovery from COVID-19 lymphopenia.

As T-cell proliferative capacity is telomere-length (TL) dependent,^7-9^ we have reasoned that short telomeres might compromise the ability to offset T-cell loss in the face of COVID-19.^10^ The ensuing deficit between T-cell loss and production would result in lymphopenia, diminish the ability to clear SARS-CoV-2, and increase the risk of severe COVID-19. This process might partially explain the increased severity and fatality of COVID-19 in older persons, because TL shortens with age.^10^

Epidemiologic studies have used leukocyte TL (LTL) as a proxy for an individual’s TL in different leukocyte lineages and somatic cells. This approach is justified, since the inter-individual variation in TL far exceeds TL differences among leukocyte lineages and somatic cells within an individual.^11-13^ These studies as well as more fundamental telomere research are based on the premise that telomeres experience attrition with somatic cell replication, a process ultimately generating short telomeres that signal the cellular machinery to stop replication. Such a signal is triggered, however, by the shortest telomeres − not the mean of the different lengths of telomeres in the nucleus.^14,15^ Almost all previous telomere studies have employed techniques that generate data based on mean TL (mTL) rather than the shortest telomeres in DNA samples. In the present study, we employed a novel method coined the Telomeres Shortest Length Assay (TeSLA) that also measures and tallies short telomeres in the TL distribution.^16^ This approach allowed us to test the relation between lymphocyte counts and short TL in patients with COVID-19, using a more direct and biologically meaningful parameter of short TL. We also quantified mTL by Southern blotting (SB),^17^ to facilitate comparison with a standard TL measurement method. Notably, we focused on older adults because of their comparatively shorter LTL^18,19^ and susceptibility to severe COVID-19.

## Methods

### Subjects

We studied 38 older participants (age 65-104 years) enrolled in the CORSER 2e branch of a SARS-Co-V-2 study in France (N° ID-RCB: 2020-A00406-33; ClinicalTrials.gov Identifier: NCT04325646). They were admitted to the Geriatric Department of the University of Nancy. Of these, 17 patients were hospitalized for COVID-19, confirmed by RT-PCR test for SARS-COV-2 in nasopharyngeal samples and clinical manifestations of COVID-19. The other 21 non-COVID-19 patients were hospitalized for different reasons; they had negative RT-PCR tests and no clinical or radiological findings consistent with COVID-19. We recorded lymphocyte count from the first blood sample collected after the RT-PCR confirmation of COVID-19 or admission blood sample in non-COVID-19 patients. Eighteen participants signed an informed consent approved by the COMITE DE PROTECTION DES PERSONNES (CPP) ILE DE France III. CPP File N °: Am8448-6-3765. Next of kin signed the informed consent for the other 20 patients who were unable to sign the document. None of the participants, including COVID-19 patients, died within 15 days after being enrolled. The COVID-19 participants do not represent all older COVID-19 patients admitted to the Geriatric Department of the University of Nancy, because some patients died before attending physicians were able to secure their informed consent.

### Measurements of TL parameters

TeSLA and SB measurements are detailed in previous communications.^16,17^ Although these two methodologies of TL measurements are correlated (see below and under Supplemental Information), they do not produce identical information. The SB method is the ‘gold standard’ of TL measurements against which the validity of all other techniques is judged, but it was designed to measure the mean length of the 92 telomeres of the q and p arms of the 23 human chromosome pairs (SB mTL). SB typically generates a weak signal for TLs below 3 kilobases (kb) and is therefore usually applied to a range of 3 to 20 kb (Figure 1). TeSLA, in contrast, generates signals for single telomeres in the TL distribution, and covers a TL range from 18 kb to below 0.8 kb (Figure 1). TeSLA measurements yield two key metrics: (a) the proportion of short telomeres (expressed in %) with different TL thresholds for which there is as yet no criterion based on ultimate end points, and (b) TeSLA mTL (in kb), which is the mean TL of the telomeres captured by TeSLA measurements.

**Figure 1.**
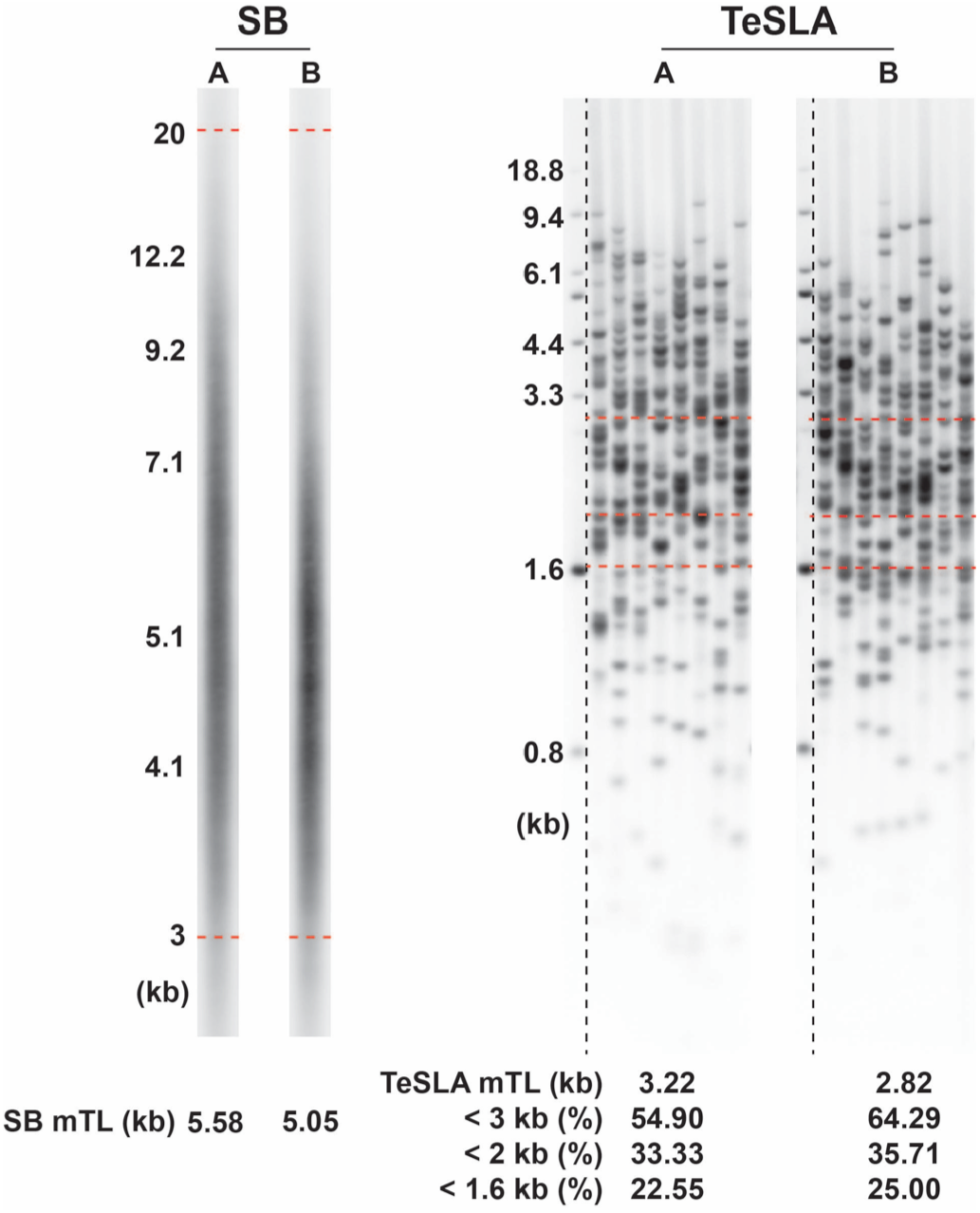
Illustrations of Southern blotting (SB) and Telomeres Shortest Length Assay (TeSLA) outputs. DNA samples are from peripheral blood mononuclear cells (PBMCs) donated by two participants in the study (designated A and B). For SB (on the left), each lane shows a single run for each DNA sample. For TeSLA (on the right), each panel comprises seven lanes and the single telomeres are shown as bands. All lanes in each panel are derived from one DNA sample. In this way, the TeSLA output generates sufficient bands required to ascertain reliability and reproducibility of the measurements. The interrupted red horizontal lines for SB display the scan limits between 20 and 3 kb. The red horizontal line for TeSLA show the 3 kb, 2 kb and 1.6 kb telomere length thresholds. Mean TL by SB (SB mTL), and mean TL by TeSLA (TeSLA mTL) and proportion of telomeres shorter < 3 kb, < 2 kb, and < 1.6kb by TeSLA are displayed at the bottoms of the lanes and panels. Molecular weight (MW) reference ladders are shown on left of the SB and TeSLA runs.

### Statistical analyses

Proportions of short telomeres, comorbidities and sex were compared using a Chi^2^-test, and correlations were tested using linear regression implemented in R.^20^ We included age and sex as fixed effects in the regression models, because age and sex could confound the relationship of TL to lymphocyte count.^18,19,21^ However, excluding age and sex from the models did not change the conclusions. The primary hypothesis we tested focused on the correlation of the lymphocyte count (dependent variable) with the proportion of telomeres < 2 kb among COVID-19 patients. We also used two alternative thresholds, i.e., < 3 kb and < 1.6 kb, to ascertain that results did not reflect an arbitrary choice of threshold (this yielded the same conclusions, results not shown). We next tested the correlation between lymphocyte count and TeSLA mTL and SB mTL. Although SB mTL and TeSLA mTL are correlated (Figure S1A), the proportions of short telomeres, e.g., < 2 kb, correlate better with TeSLA mTL (Figure S1B) than with SB mTL (Figure S1C). Finally, we performed these analyses in non-COVID-19 patients, and compared correlations with those among COVID-19 patients.

## Results

General characteristics of COVID-19 and non-COVID-19 subjects are displayed in Table 1. TL parameters by TeSLA and SB for the two patient groups are displayed in Table S1.

**Table 1.** General characteristics

|  | COVID-19<br>No<br>(N=21) | COVID-19<br>Yes<br>(N=17) | P |
| --- | --- | --- | --- |
| **Characteristics** |  |  |  |
| Women – N (%) | 12 (57) | 9 (53) | 0.79 |
| Age (years) | 87 ± 9 | 87 ± 8 | 0.96 |
| BMI (kg/m<sup>2</sup>) | 24.7 ± 5.6 | 26.2 ± 6.0 | 0.62 |
| **Comorbidities –N (%)** |  |  |  |
| Cardiovascular disease | 11 (52) | 11 (65) | 0.44 |
| Pulmonary disease | 3 (14) | 3 (18) | 0.78 |
| Cancer | 4 (19) | 3 (18) | 0.91 |
| Neurological disease | 8 (38) | 9 (53) | 0.36 |
| Diabetes mellitus | 7 (33) | 2 (12) | 0.12 |
| Hypertension | 15 (71) | 13 (76) | 0.73 |
| **Medications** |  |  |  |
| Number of chronic medications | 6.6 ± 3.2 | 6.4 ± 3.7 | 0.58 |
| **Blood and Kidney Parameters** |  |  |  |
| Lymphocytes (10<sup>9</sup>/L) | 1.50 ± 0.50 | 0.91 ± 0.42 | <0.001 |
| Leucocytes (10<sup>9</sup>/L) | 6.49 ± 2.06 | 5.72 ± 2.21 | 0.27 |
| Hemoglobin (g/dL) | 12.0 ± 1.8 | 11.5 ± 1.8 | 0.41 |
| CRP (mg/L) | 17.9 ± 13.5 | 63.2 ± 53.2 | <0.001 |
| Creat.clear. (ml/min/1.73m<sup>2</sup>)* | 79.1 ± 21.1 | 74.1 ± 25.8 | 0.30 |

BMI, body mass index; C reactive protein, CRP; \* Modification of Diet in Renal Disease method; data are presented as number or mean ± standard deviation as indicated.

Lymphocyte count (10^9^/L) was significantly lower in COVID-19 patients compared to non-COVID-19 patients (difference: -0.59 ± 0.15; P < 0.001), and this difference remained virtually unchanged (difference: -0.57 ± 0.098; P < 0.001) when controlling for sex and age.

Among COVID-19 patients, lymphocyte count showed significant inverse correlations with the proportion of short telomeres (P = 0.005; Figure 2;) and this relationship remained unchanged after adjusting for age and sex (P = 0.005, Table S2). Lymphocyte count in non-COVID-19 patients showed no significant correlation with the proportion of short telomeres (Figure 2, Table S2). Consistent with these findings, lymphocyte count was positively correlated with TeSLA mTL (P = 0.03; Figure S2A; Table S2). This correlation was not significant among non-COVID-19 patients (Figure S2B; Table S2). Patterns for COVID-19 patients and non-COVID-19 patients differed significantly with regard to the proportion of short telomeres (interaction COVID-19 status * proportion short telomeres: P = 0.0013; P <0.02 for interaction with TeSLA mTL). Finally, For COVID-19 patients, the correlation of lymphocyte count with SB mTL showed a pattern similar to the TeSLA data (Figure 3), but did not reach statistical significance (Table S3).

**Figure 2.**
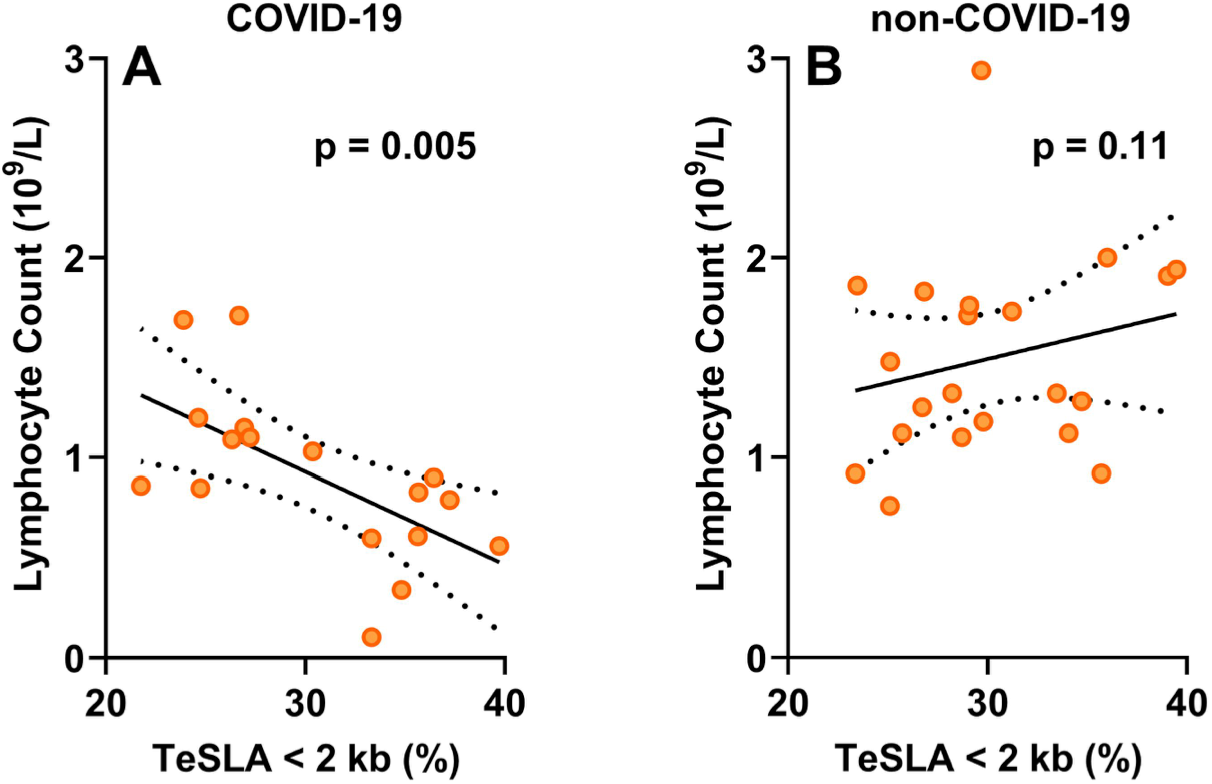
Lymphocyte count in relation to proportion of telomeres <2 kb in COVID-19 patients (A) and non-COVID-19 patients (B). See table S2 for the statistical models.

**Figure 3.**
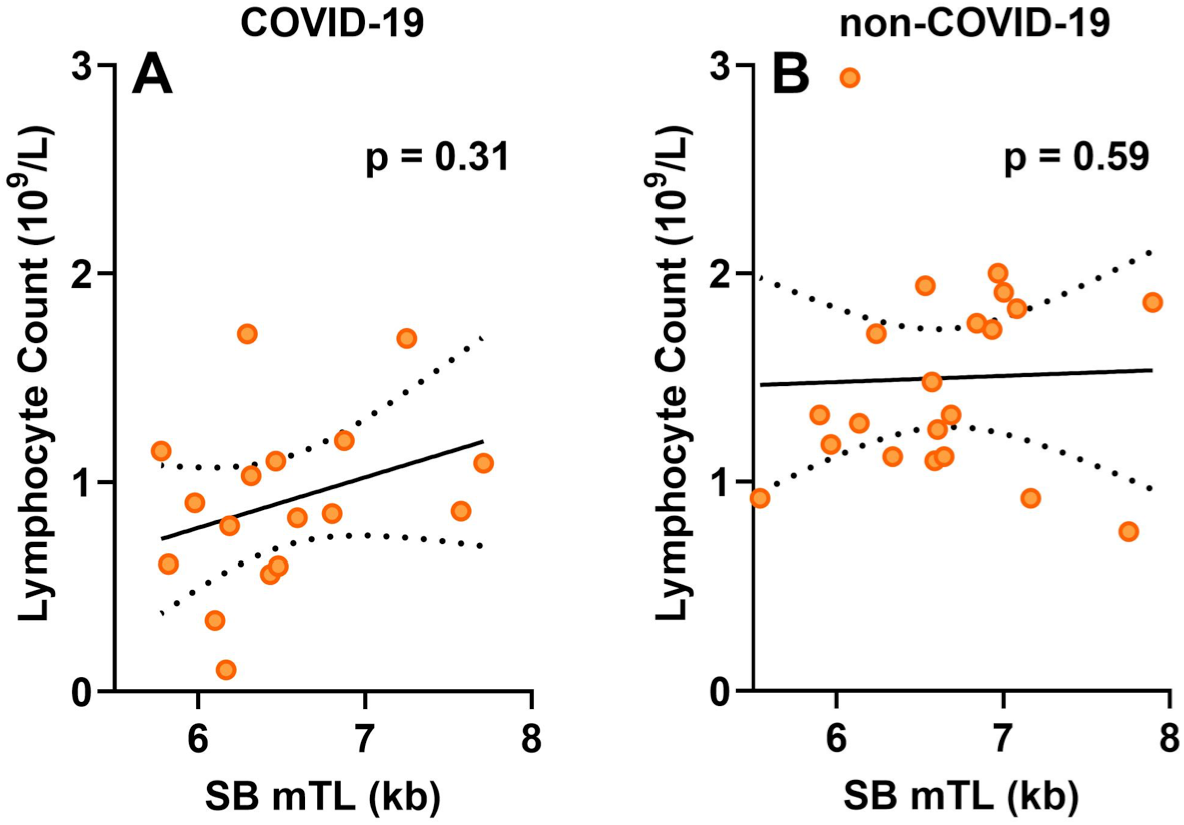
Lymphocyte count in relation to the Southern blotting mean telomere length (SB mTL) in COVID-19 patients (A) and non-COVID-19 patients (B). See table S3 for the statistical models.

## Discussion

Three well-established findings build the foundation for our model. First, the plummeting number of T-cells is the principal cause of COVID-19 lymphopenia.^1-4^ Second, TL parameters in PBMCs largely reflect TL parameters in all leukocyte lineages, including T-cells.^12^ Third, the shortest telomeres in the TL distribution− which can be detected by TeSLA− are better indicators than mTL of telomere-related biological endpoints.^14,15^

The T-cell blood pool is a component of the T-cell hierarchy, whose structure and composition are complex and change with age.^22,23^ For simplicity, we consider the T-cell blood pool as a homogeneous entity (Figure 4). In this simplified scenario, the size of the T-cell blood pool reflects the balance between T-cell depletion - due to senescence/death and sequestration out of the circulation – and T-cell repletion through proliferation. As T-cell proliferation is TL-dependent,^7-9^ individuals with extremely short telomeres because of rare detrimental mutations are unable to maintain their T-cell blood pool.^24^ In the general population, however, TL variation shows no relation with lymphocyte count ^25^, presumably because of the low turnover of T-cells ^26^ (Figure 4A). This might not apply in the face of COVID-19, since rapid T-cell proliferation is probably critical for offsetting the development of, and recovery from, COVID-19 lymphopenia. Accordingly, we hypothesized that when infected with SARS-CoV-2, persons with short telomeres lag in their T-cell proliferative response, resulting in a deficit in their T-cell blood pool (Figure 4B).^10^ Our results support this hypothesis. They also suggest that COVID-19 exposes the limited TL-dependent replicative reserves of T cells in older persons with short telomeres.

**Figure 4.**
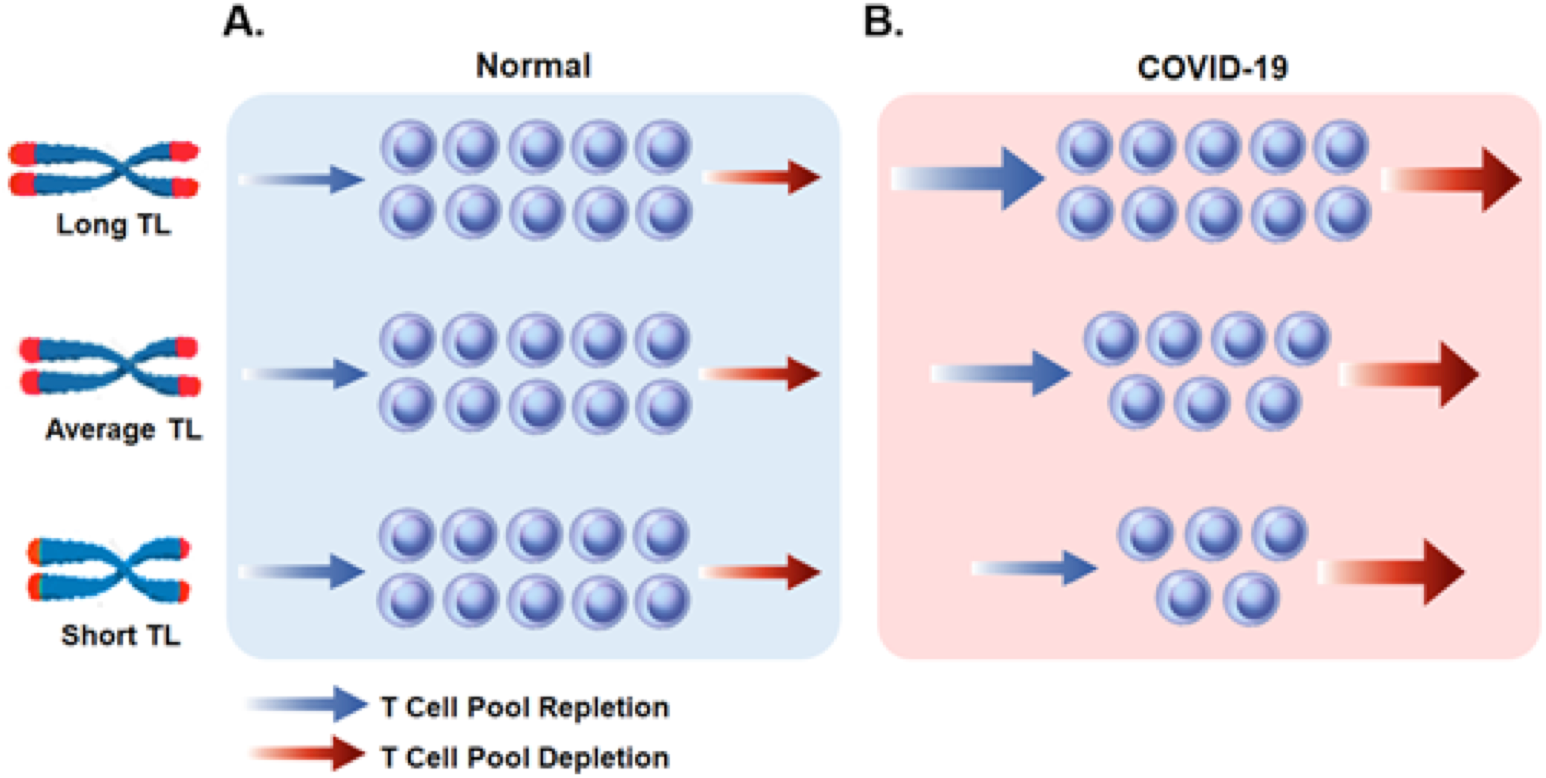
Depletion and repletion of the T cell blood pool under normal condition and during COVID-19. Under normal, ‘steady-state’ condition (A), persons with shorter telomeres maintain their T cell blood pool. However, in the face of COVID-19 (B), persons with shorter telomeres might lag in their ability to increase T cell production (increase in the pace of the T Cell Pool Repletion− arrow) to match increased T cell loss due to the infection (Increase in the pace of T Cell Pool Depletion− arrow). This non-steady condition would result in shrinking of the T cell pool in proportion to the shorter telomeres. Telomeres are displayed as red caps at the ends of the chromosomes.

Data from children with COVID-19 further support our model. Children, who are otherwise healthy, typically show a mild clinical course when infected with SARS-CoV-2. Whereas lymphopenia is a major prognostic feature of COVID-19 in adults, it is a minor aspect of little prognostic value in children with COVID-19.^27-30^ The average LTL in children is ∼2 kb longer than in adults;^19,21^ hence children have the capacity to accelerate T-cell production to offset even a drastic T-cell loss in SARS-CoV-2 infection. However, given the wide inter-individual TL variation in the general population, TL of some young adults in their 30s and 40s is as short as TL in older persons in their 70s and 80s. ^18,19^ These young adults, we propose, may be susceptible to COVID-19 lymphopenia despite their young age.

We acknowledge limitations of this study. The cross-sectional data we provide do not prove causality. Reverse causation, however, seems unlikely; a low lymphocyte count would not rapidly shorten TL in COVID patients. The detection of the inverse correlation between lymphocyte count and the proportion of very short telomeres required use of TeSLA, a novel and expensive approach available in few laboratories worldwide. Nonetheless, the correlation between lymphocyte count and SB mTL showed a pattern similar to that generated by TeSLA mTL. As TeSLA and SB results are correlated, we anticipate that the TL-COVID-19 connection we highlight might also be captured using SB measurements in larger cohorts. In addition, the clinical picture and outcomes of COVID-19 are complex and multi-factorial and TL dynamics of hematopoietic cells are only a component of the disease. Finally, we studied a small cohort of older persons of European ancestry, all surviving the disease for at least 15 days. This most vulnerable group to severe COVID-19 is not representative of the general population, but studying its members has provided the optimal setting to examine the TL-lymphopenia connection in this disease.^10^

In conclusion, in a small study of older adults, we tested a model that links lymphopenia in older COVID-19 patients with PBMC TL parameters. Our findings show that comparatively short PBMC telomeres, as expressed in the shortest telomeres of the TL distribution, are associated with diminished lymphocyte count in older persons infected with SARS-CoV-2. Our findings point the way for larger investigations in the general adult population, focusing on T-cell TL parameters and their role in COVID-19 lymphopenia.

## Data Availability

All data are available upon request to the corresponding author

## Supplementary Information

**Figure S1.**
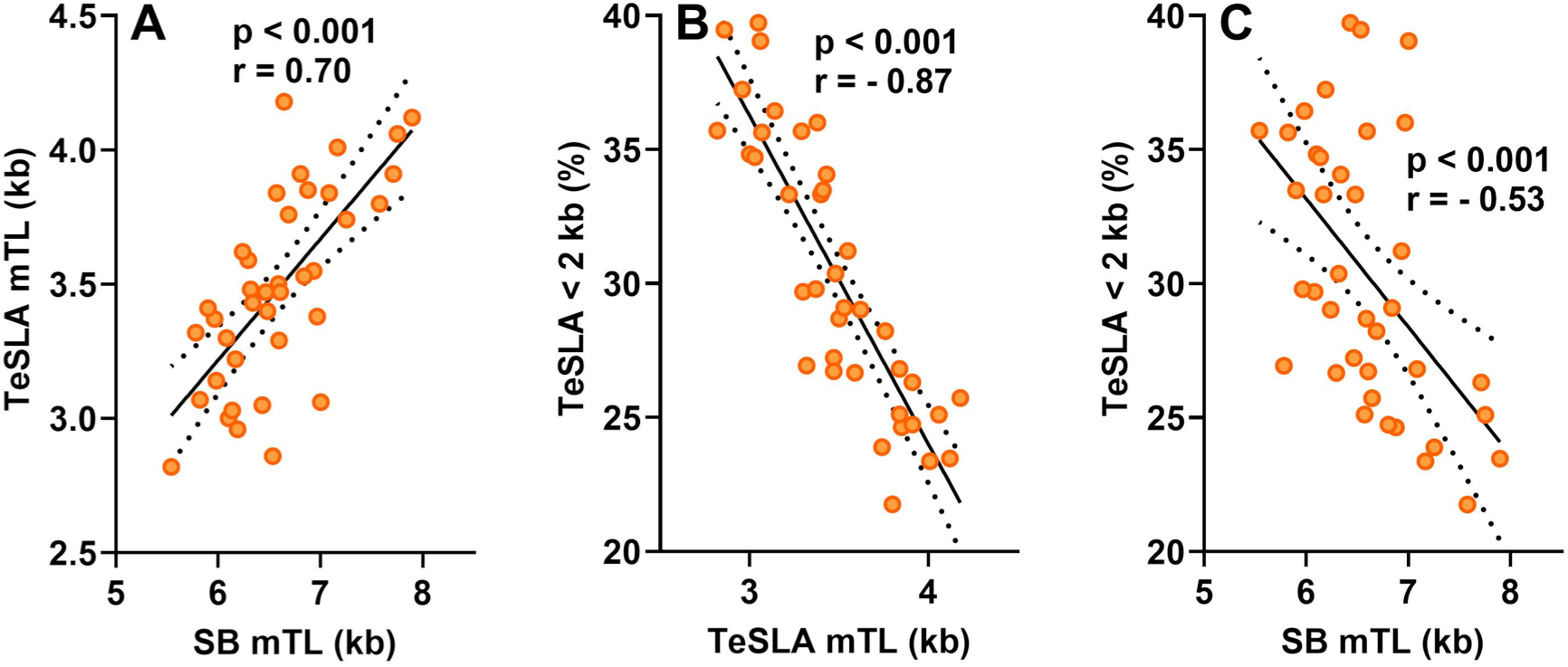
Comparison of the two methods to measure telomeres, Southern blot and Telomeres Shortest Length Assay (TeSLA). SB mTL, mean telomere length using Southern blots; TeSLA mTL, mean telomere length using TeSLA; TeSLA <2 kb (%) is the percentage of telomeres shorter than 2 kb. Data are for telomere parameters from all study participants.

**Figure S2.**
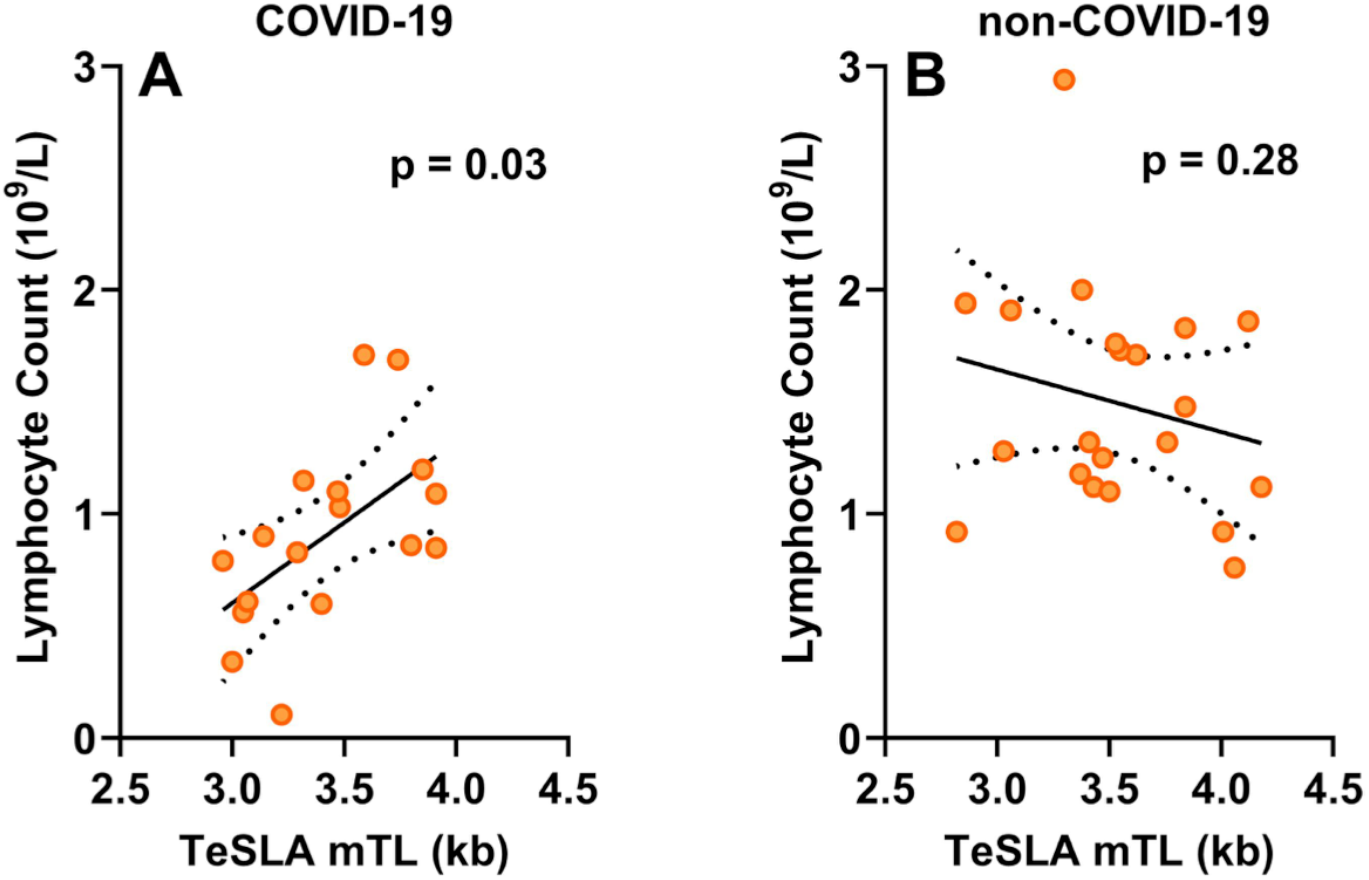
Lymphocyte count in relation to the mean of the short telomeres as generated by TeSLA (TeSLA mTL) in COVID-19 patients (A) and non-COVID-19 patients (B).

**Table S1.** Leukocyte telomere parameters

|  | **COVID-19**<br>**No**<br>**(N=21)** | **COVID-19**<br>**Yes**<br>**(N=17)** | **P** |
| --- | --- | --- | --- |
| TeSLA mTL (kb) | 3.53 ± 0.39 | 3.42 ± 0.33 | 0.37 |
| TeSLA TL <1.6 kb (%) | 21.4 ± 4.0 | 21.5 ± 5.0 | 0.95 |
| TeSLA TL <2 kb (%) | 30.2 ± 4.9 | 30.5 ± 5.6 | 0.95 |
| TeSLA TL <3 kb (%) | 51.0 ± 6.4 | 52.6 ± 6.2 | 0.35 |
| SB mTL (kb) | 6.64 ± 0.58 | 6.52 ± 0.57 | 0.35 |

TeSLA, Telomere Shortest Length Assay; SB, Southern blotting; TL, telomeres length. Data are presented as mean ± standard deviation.

**Table S2.** Multiple regression analysis of the relation of lymphocyte count with age, sex and telomere length parameters measured using TeSLA in COVID-19 patients (n=17, Tables A, B) and non-COVID-19 patients (n=21, Tables C, D). Sub-tables used different telomere length parameters as indicated in the Table headers. Estimates of sex effect indicate how males differ from females.

**A. COVID-19 patients. Proportion of telomeres shorter than 2.0 kb. $R^2 = 0.68$**
| Variable | Estimate ± SE | *t* | P |
| --- | --- | --- | --- |
| Intercept | 4.04 ± 0.79 | 5.10 | 0.0002 |
| Telomeres <2 kb (%) | -0.042 ± 0.013 | 3.32 | 0.005 |
| Age (years) | -0.019 ± 0.009 | 2.03 | 0.06 |
| Sex | -0.42 ± 0.13 | 3.22 | 0.007 |

**B. COVID-19 patients. TeSLA mTL. $R^2 = 0.59$**
| Variable | Estimate ± SE | *T* | P |
| --- | --- | --- | --- |
| Intercept | 1.13 ± 1.39 | 0.81 | 0.43 |
| mTL (kb) | 0.56 ± 0.23 | 2.39 | 0.03 |
| Age (years) | -0.23 ± 0.010 | 2.17 | 0.05 |
| Sex | -0.37 ± 0.15 | 2.47 | 0.03 |

**C. Non-COVID-19 patients. Proportion of telomeres shorter than 2.0 kb. $R^2 = 0.45$ .**
| Variable | Estimate ± SE | *t* | P |
| --- | --- | --- | --- |
| Intercept | 0.17 ± 1.06 | 0.16 | 0.87 |
| Telomere <2 kb (%) | 0.037 ± 0.022 | 1.68 | 0.11 |
| Age (years) | 0.005 ± 0.012 | 0.44 | 0.67 |
| Sex | -0.53 ± 0.21 | 2.58 | 0.02 |

**D. Non-COVID-19 patients. TeSLA mTL. $R^2 = 0.27$**
| Variable | Estimate ± SE | *t* | P |
| --- | --- | --- | --- |
| Intercept | 2.74 ± 1.96 | 1.40 | 0.18 |
| mTL (kb) | -0.34 ± 0.31 | 1.11 | 0.28 |
| Age (years) | 0.002 ± 0.013 | 0.14 | 0.89 |
| Sex | -0.47 ± 0.21 | 2.24 | 0.04 |

**Table S3.** Multiple regression analysis of the relation between lymphocyte count with age, sex and mTL measured by Southern blotting in A. COVID-19 patients (n=17) and B. non-COVID-19 patients (n=21).

**A. COVID-19 patients. $R^2 = 0.34$**
| **Variable** | **Estimate ± SE** | ***t*** | **P** |
| --- | --- | --- | --- |
| **Intercept** | 2.52 ± 1.50 | 1.6 | 0.12 |
| **mTL (kb)** | 0.165 ± 0.155 | 1.07 | 0.31 |
| **Age (years)** | -0.0290 ± 0.011 | 2.52 | 0.03 |
| **Sex** | -0.362 ± 0.176 | 2.06 | 0.06 |

**B. Non-COVID-19 patients. $R^2 = 0.48$**
| **Variable** | **Estimate ± se** | ***t*** | **P** |
| --- | --- | --- | --- |
| **Intercept** | -0.135 ± 2.121 | 0.06 | 0.95 |
| **mTL (kb)** | 0.112 ± 0.204 | 0.55 | 0.59 |
| **Age (years)** | 0.012 ± 0.013 | 0.93 | 0.36 |
| **Sex** | -0.441 ± 0.212 | 2.08 | 0.05 |

## Notes

### Competing Interest Statement

The authors have declared no competing interest.

### Clinical Trial

NCT04325646

### Funding Statement

This study has been supported by the French National Research Agency (ANR), Translationnelle: ID RCB: 2014-A00298-39: 2014-2017 and partially supported by the French PIA project Lorraine Universite d Excellence, reference ANR-15-IDEX-04-LUE and the Investments for the Future program under grant agreement ANR-15-RHU-0004.
AA research is supported by National Institutes of Health grants R01HL134840, U01AG066529, and Norwegian Institute of Public Health grants 262700 and 262043.

### Author Declarations

ETHICS APPROVAL STATEMENT This study was approved by the COMITE DE PROTECTION DES PERSONNES, ILE DE France III. CPP File Number: Am8448-6-3765, and by the Rutgers University, New Jersey Medical School IRB, Pro2020000669.

